# Correlation of coagulation parameters with clinical outcomes in Coronavirus-19 affected minorities in United States: Observational cohort

**DOI:** 10.1101/2020.05.01.20087932

**Authors:** Morayma Reyes Gil, Jesus D. Gonzalez-Lugo, Shafia Rahman, Mohammad Barouqa, James Szymanski, Kenji Ikemura, Yungtai Lo, Henny H Billett

## Abstract

**Importance:** COVID-19 has caused a worldwide illness and New York has become the epicenter of COVID-19 in the United States. Currently Bronx has the highest prevalence per capita in New York.

**Objective:** To investigate the coagulopathic presentation of COVID and its natural course and to investigate whether hematologic and coagulation parameters can be used to assess illness severity and death.

**Design:** Retrospective case study of positive COVID inpatients between 3/20/2020-3/31/2020.

**Setting:** Montefiore Health System main hospital, Moses, a large tertiary care center in the Bronx.

**Participants:** Adult inpatients with positive COVID tests hospitalized at MHS.

**Exposure (for observational studies):** Datasets of participants were queried for physiological, demographic (age, sex, socioeconomic status and self-reported race and/or ethnicity) and laboratory data.

**Main Outcome and Measures:** Relationship and predictive value of measured parameters to mortality and illness severity.

**Results:** Of the 217 in this case review, 70 died during hospitalization while 147 were discharged home. Only the admission PT and first D-Dimer could very significantly differentiate those who were discharged alive and those who died. Logistic regression analysis shows increased odds ratio for mortality by first D-Dimer within 48 hrs. of admission. The optimal cut-point for the initial D-Dimer to predict mortality was found to be 1.65 μg/mL

**Conclusions:** We describe here a comprehensive assessment of hematologic and coagulation parameters in COVID and examine the relationship of these to mortality. We demonstrate that both initial and maximum D-Dimer values are biomarkers that can be used for survival assessments.

## BACKGROUND

COVID-19 is a heterogenous disease caused by 2019-nCoV/SARS-CoV-2 virus, a new member of the corona virus. Clinical manifestations vary from an asymptomatic illness in some to rapid death in others (1–4). COVID-19 has caused a worldwide illness, causing WHO to declare this a pandemic on January 30^th^ 2020.(5) The United States is currently the global hot spot with a fatality rate ranging from 10% to 27% among adults aged more than 85 years, 3% to 11% among adults of age group 65- 84 years and 1% to 3% among age group 55- 64 years;(5) New York is the epicenter of COVID-19 in the United States and the Bronx currently has the highest prevalence per capita in New York (6, 7).

Our hospital is a large tertiary care center and is the primary medical system for the Bronx. Faced with a human crisis of unprecedented proportions, we decided to try and understand this disease, not only the presentation but the potential correlations with illness severity and death and the natural course of both complicated and less complicated disease. The primary objective of this study is to examine demographic and laboratory data, as well as changes in laboratory data, and to determine the relation between these laboratory parameters, in particular tests of coagulation, to illness severity. In addition, we wanted to examine whether the Bronx, with one of the largest minority populations in the country, as well as one of the poorest, was at greater risk than other areas in the country.

Early on in the course of taking care of these patients, it became obvious that there was a significant prothrombotic diathesis to this disease. The coagulation analyzers required cleansing several times a day rather than once a week; the dialysis lines were clotting up; young patients with normal coronary arteries were having myocardial infarctions and others were abruptly dying from what might be assumed to be pulmonary emboli. Coagulopathy and D-Dimer elevations are reported in 3.75- 68.0% of the COVID-19 patients (8). Studies from patients in Wuhan showed an association of high D-Dimer, a marker for thrombosis, with mortality (9). Herein we studied if D-Dimer and other parameters correlated with mortality in the Bronx population.

## METHODS

### STUDY DESIGN

#### Data Gathering and Variables

In our retrospective, single-center study, we included confirmed COVID-19 cases in Montefiore Medical Center/University Hospital for Albert Einstein College of Medicine, Moses Campus, who were hospitalized and had routine coagulation tests done between March 20^th^ to March 31^st^ 2020. For those samples without an ordered D-dimer, D-Dimer was performed alongside with prothrombin time (PT) as part of this study. All cases were established with reverse-transcriptase–polymerase-chain-reaction real-time (RT PCR) assay of the nasal and the pharyngeal swabs. We excluded the patients younger than 18 years of age and those admitted for COVID-19 illness despite negative test results or if any of the data was missing. The study was approved by the Albert Einstein College of Medicine Institutional Review Board. The medical record of the patients was reviewed to obtain the epidemiological, demographic, clinical and laboratory data.

These variables included demographic attributes (age, sex and self-reported race and/or ethnicity) and baseline comorbidities (body mass index, previous history of hypertension, diabetes, kidney, pulmonary, liver, autoimmune, cancer, or sickle cell, disease on presentation). We gathered data on vital signs and laboratory values upon presentation to the hospital and initial laboratory values. D-dimer in the first 48 hrs. of presentation to ED and/or hospital admission was noted. The laboratory assessments consisted of a complete blood count, blood chemical analysis, coagulation testing, assessment of liver and renal function, and measures of electrolytes, prothrombin time (PT), partial thromboplastin time (PTT), D-Dimer, fibrinogen, C-reactive protein, procalcitonin. The management and clinical outcomes were followed up to April 20, 2020. We assessed for interventions and time to interventions including ICU admission, intubation, and anticoagulation. Discharge from hospital and mortality were assessed.

The earliest symptoms were categorically defined: New onset cough, dyspnea and diarrhea. Oxygen saturation on room air, intubation and dialysis requirements were documented. Maximum and when appropriate minimum, values were noted and day of these values from admission date were noted. Daily levels of parameters under particular scrutiny were noted whenever possible. Obesity was defined as BMI more than 30. For history of prior comorbidities, cardiac comorbidity was defined as the presence of hypertension, coronary artery disease, or heart failure. Cancer was defined as malignancy with active treatment or diagnosed within the last five years. Imaging study was defined as the chest imaging done on presentation to the emergency department. All patients who were still inpatients as of that date were excluded in the data analysis.

#### Laboratory Testing

All cases were established with reverse-transcriptase–polymerase-chain-reaction real-time (RT PCR) assay of the nasal and the pharyngeal swabs Coagulation tests (prothrombin time, D-dimer, partial thromboplastin time and fibrinogen) were performed by STA-R Max instruments. STA Liatest LIA D-dimer assay was performed as per manufacturer recommendations and reported as FEU μg/mL with a cut off of <0.5 ug/ml to rule out PE. Complete blood counts were performed by Sysmex XN9000. Chemistry assays were performed by Roche instrumentation and reagents as per manufacturer recommendations.

#### Statistical Methods

Data analysis was performed using R software, version 3.6.2. Differences in demographic, clinical variables and laboratory assessments between patients died in hospital and patients discharged alive were compared using chi-square tests, or Fisher’s exact tests for categorical variables and two-sample Student *t* tests, or the Mann-Whitney U test for continuous variables. Logistic regression was carried out to examine the relationship between the factors and lab parameters under examination and in-hospital mortality. The receiver operating characteristic curves (ROC) were used to assess performance of D-Dimer in the first 48 hours on predicting in-hospital mortality adjusted for age, BMI and sex.

### RESULTS

#### Study Population

*Admission and Mortality data*. Of the 217 patients who tested positive for COVID, 70 (32%) patients died during hospitalization while 147 (68%) were discharged alive. Similar to other reports, patients that succumbed to death were significantly older than survivors with a mean (± SD) of 68.7 ± 12.4 vs. 57.7 ± 15.6 years (p <0.001). Analysis of physiological parameters, comorbidities, racial and other demographics are included in Table 1. The only clinical and physiological parameter on admission that showed statistical significance between non-survivors vs. survivors was the oxygen saturation <93% on RA (66% vs. 24%, p <0.0001). We could not detect a statistical significance for gender or race between survivors vs. non-survivors in this population.

**Table 1.** Demographics and clinical features of COVID positive patients at admission.

| Demographics/ Clinical Features | Died<br>n=70 | Discharged<br>n=147 | P value |
| --- | --- | --- | --- |
| Age, yrs [mean (SD)] | 68.71 (12.44) | 57.71 (15.56) | <0.001 |
| Gender |  |  |  |
| - Female, n (%) | 23 (32.9) | 68(46.3) | 0.085 |
| - Male, n (%) | 47 (67.1) | 79 (53.7) |  |
| Comorbidity |  |  |  |
| - Hypertension, n (%) | 52 (74.3) | 90 (61.2) | 0.068 |
| - Diabetes, n (%) | 32 (45.7) | 49 (33.3) | 0.11 |
| - Chronic Kidney Disease, n (%) | 14 (20.0) | 23 (15.6) | 0.44 |
| - Pulmonary Disease, n (%) | 16 (22.9) | 35(23.8) | 1.0 |
| - Liver Disease, n (%) | 4 (5.7) | 6 (4.1) | 0.73 |
| - Autoimmune Disease, n (%) | 4 (5.7) | 8 (5.4) | 1.0 |
| - Cancer, n (%) | 9 (12.9) | 8 (5.4) | 0.10 |
| - Sickel Cell Disease, n (%) | 1 (1.4) | 1 (0.7) | 0.54 |
| Race |  |  |  |
| - Black, n (%) | 32 (46) | 53 (36) | 0.36 |
| - White, n (%) | 9 (13) | 29 (20) |  |
| - Asian, n (%) | 2 (3) | 2 (1) |  |
| - Other/Declined, n (%) | 27 (38) | 63 (43) |  |
| Ethnicity |  |  |  |
| - Hispanic, n (%) | 19 (27) | 53 (36) | .09 |
| - Non- Hispanic, n (%) | 47 (67) | 77 (52) |  |
| - Other/Declined, n (%) | 4 (6) | 17 (12) |  |
| BMI (median (IQR)) | 29.02 (26.00, 33.12) | 29.54 (25.94, 33.53) | 0.79 |
| Presentation |  |  |  |
| - Fever, n (%) | 51 (72.9) | 100 (68) | 0.57 |
| - Cough, n (%) | 51 (72.9) | 103 (70.1) | 0.79 |
| - SOB, n (%) | 52(74.3) | 95(64.6) | 0.17 |
| - Diarrhea , n (%) | 9 (12.9) | 33 (22.4) | 0.10 |
| - Infiltrate on initial X-ray, n (%) | 65 (93) | 132 (90) | 0.62 |
| - O2 sat <93% on RA, n (%) | 46 (66) | 35 (24) | <0.0001 |

#### Clinical Characteristics

As shown in Table 2, intubation, cardiac arrest, dialysis requirement or significant liver disease during hospitalization was associated with decreased survival. Both the number of patients that required intensive care and the length of stay in intensive care were higher in non-survivors than survivors. The overall length of stay for survivors discharged home was significantly shorter for patients that died during hospitalization.

**Table 2:** Laboratory and clinical course of COVID positive patients during hospitalization.

|  | Died<br>n=70 | Discharged<br>n= 147 | P |
| --- | --- | --- | --- |
| **Lab Test** |  |  |  |
| Absolute Neutrophilic Count, x10<sup>9</sup>/L ( median (IQR)) |  |  |  |
| - Initial (median (IQR)) | 4.40 (3.25, 6.60) | 5.35 (3.42, 7.58) | 0.04 |
| - Minimum (median (IQR)) | 4.8 (3.00, 6.65) | 3.0 (2.20, 4.00) | <0.0001 |
| - Hospital days to minimum | 1.83 (3.33) | 4.11 (5.11) | <0.0001 |
| - Maximum (median (IQR)) | 12.65 (9.75,16.30) | 5.85 (4.10, 8.50) | <0.0001 |
| - Hospital days to maximum | 7.27(4.68) | 4.14 (10.53) | 0.24 |
| Absolute Lymphocytic Count, x10<sup>9</sup>/L (median (IQR)) |  |  |  |
| - Initial | 0.90 (0.70, 1.20) | 0.90 (0.70, 1.20) | 0.80 |
| - Minimum | 0.60 (0.40-0.80) | 0.80 (0.60- 1.10) | <0.0001 |
| - Hospital days to minimum | 4.23 (4.04) | 2.29 (3.33) | 0.43 |
| Hemoglobin, g/dl (mean (SD)) |  |  |  |
| - First | 12.77 (2.19) | 12.69 (2.51) | 0.80 |
| - Minimum | 9.8 (2.8) | 11.1 (2.5) | 0.0014 |
| - Hospital days to maximum | 6.77 (5.34) | 4.73 (4.72) | 0.57 |
| Platelets, x10<sup>9</sup>/L ((mean (SD)) |  |  |  |
| - Initial | 206.6 (78.6) | 199.3 (71.8) | 0.515 |
| - Minimum | 154 (65.1) | 185 (71.5) | 0.0033 |
| - Hospital days to minimum | 2.87 (3.98) | 1.80 (2.96) | 0.96 |
| Procalcitonin, ng/ml (mean (SD)) |  |  |  |
| - Initial | 3.31 (9.26) | 0.63 (1.40) | 0.052 |
| - Maximum | 10.01 (15.78) | 0.97 (2.50) | 0.00026 |
| Initial AST (median (IQR)) | 51.00 [53.75] | 38.0 (34.5) | 0.018 |
| Initial ALT (median (IQR)) | 30.0 (23.50) | 28.0 (25.5) | 0.58 |
| Initial Creatinine | 1.88 (1.54) | 1.66 (1.93) | 0.38 |
| **Clinical Course** |  |  |  |
| Cardiac arrest (%) | 33 (47.1) | 1 (0.7) | <0.001 |
| Dialysis required (%) | 37 (52.9) | 9 (6.1) | <0.001 |
| LFT> 2.5x ULN at any time | 13.0 (18.6%) | 13.0 (8.8%) | 0.039 |
| Intubation required (%) | 45(64.3) | 10 (6.8) | <0.001 |
| Days to intubation (median (IQR)) | 0 (0-0) | 0 (0-2) | <0.00001 |
| Intubated days (median (IQR)) | 0 (0-0) | 3 (0-8) | <0.00001 |
| ICU LOS, days (median (IQR)) | 0 (0-0) | 0 (0-5) | <0.00001 |
| LOS, days (median (IQR)) | 9.5 (5.8-14) | 6.0 (5.6) | 0.0006 |

#### Laboratory data

These are shown in Table 2 and 3. The only significantly different admission lab tests between deceased vs. survivors were: PT and D-Dimer within first 48 hrs. (Table 3). D-Dimer results within first 48 hrs. of admission were missing in 44% non-survivors and 33% of survivors. The maximum PT, PTT, D-Dimer, procalcitonin, absolute neutrophil count (ANC) were statistically significantly higher in non-survivors vs. survivors. Likewise, the minimum absolute lymphocyte count (ALC), ANC, hemoglobin and platelet count were statistically significantly lower in non-survivors compared to survivors.

**Table 3:** Coagulation parameters and anticoagulation treatments during hospitalization.

|  | **Died**<br>**n=70** | **Discharged**<br>**N=147** | **p** |
| --- | --- | --- | --- |
| **Coagulation Profile** |  |  |  |
| **PT sec (Median (IQR))** |  |  |  |
| - Initial | 13.8 (13.3, 14.5) | 14.5 (13.6, 16.1) | 0.004 |
| - Maximum | 16.3 (15.2, 19.0) | 14.6 (13.7, 15.6) | <0.0001 |
| - Hospital days to maximum (mean (SD)) | 5.01 (4.97) | 3.39 (3.98) | 1.00 |
| **PTT sec (Median (IQR))** |  |  |  |
| - Initial | 32.5 (29.7, 35.8) | 34.0 (30.7, 37.8) | 0.098 |
| - Maximum | 39.4 (33.1, 48.8) | 34.1 (30.9, 39.6) | 0.0002 |
| - Hospital days to maximum (mean (SD)) | 4.31 (4.79) | 2.30 (4.66) | 0.99 |
| **D-Dimer ug/ml FEU (mean (SD))** |  |  |  |
| - Within first 48 hours (mean (SD)) | 4.46 (6.08) | 1.73 (1.96) | <0.001 |
| - Maximum (mean (SD)) | 7.9 (7.3) | 2.6 (3.5) | <0.0001 |
| - Hospital days to maximum | 6.29 (5.87) | 3.92 (4.61) | 0.99 |
| **Fibrinogen mg/dl (mean (SD))** |  |  |  |
| - Initial | 645.61 (253.46) | 675.41 (296.88) | 0.719 |
| - Maximum | 703.55 (259.01) | 728.68 (318.82) | 0.78 |
| - Hospital days to maximum (mean (SD)) | 6.95 (6.03) | 5.95 (5.62) | 0.57 |
| **Anticoagulation during hospitalization** |  |  |  |
| No Anticoagulation, n (%) | 13 (18.6%) | 26 (17.7%) | 0.71 |
| Prophylactic Anticoagulation, n (%) | 44 (62.9%) | 103 (70.1%) | .30 |
| LMWH | 37 | 88 |  |
| DOAC (apixaban) | 0 | 1 |  |
| Unfractionated Heparin | 6 | 14 |  |
| Warfarin | 1 | 0 |  |
| Bivalirudin | 0 | 0 |  |
| Prior AC, n (%) | 9 (12.9) | 12 (8.2) | 0.27 |
| Therapeutic Anticoagulation, n (%) | 13 (18.6%) | 19 (12.9%) | .30 |
| LMWH | 0 | 3 |  |
| DOAC (apixaban) | 5 | 12 |  |
| Unfractionated Heparin | 5 | 2 |  |
| Warfarin | 1 | 2 |  |
| Bivalirudin | 2 | 0 |  |

On multivariable logistic regression older age (OR = 1.08, 95% C.I. 1.04 – 1.12; p < 0.001) and increased first 48 hour D-Dimer (OR = 1.21, 95% C.I. 1.06 – 1.38; p = 0.05) were associated with increased odds for mortality (Figure 1).

**Figure 1.**
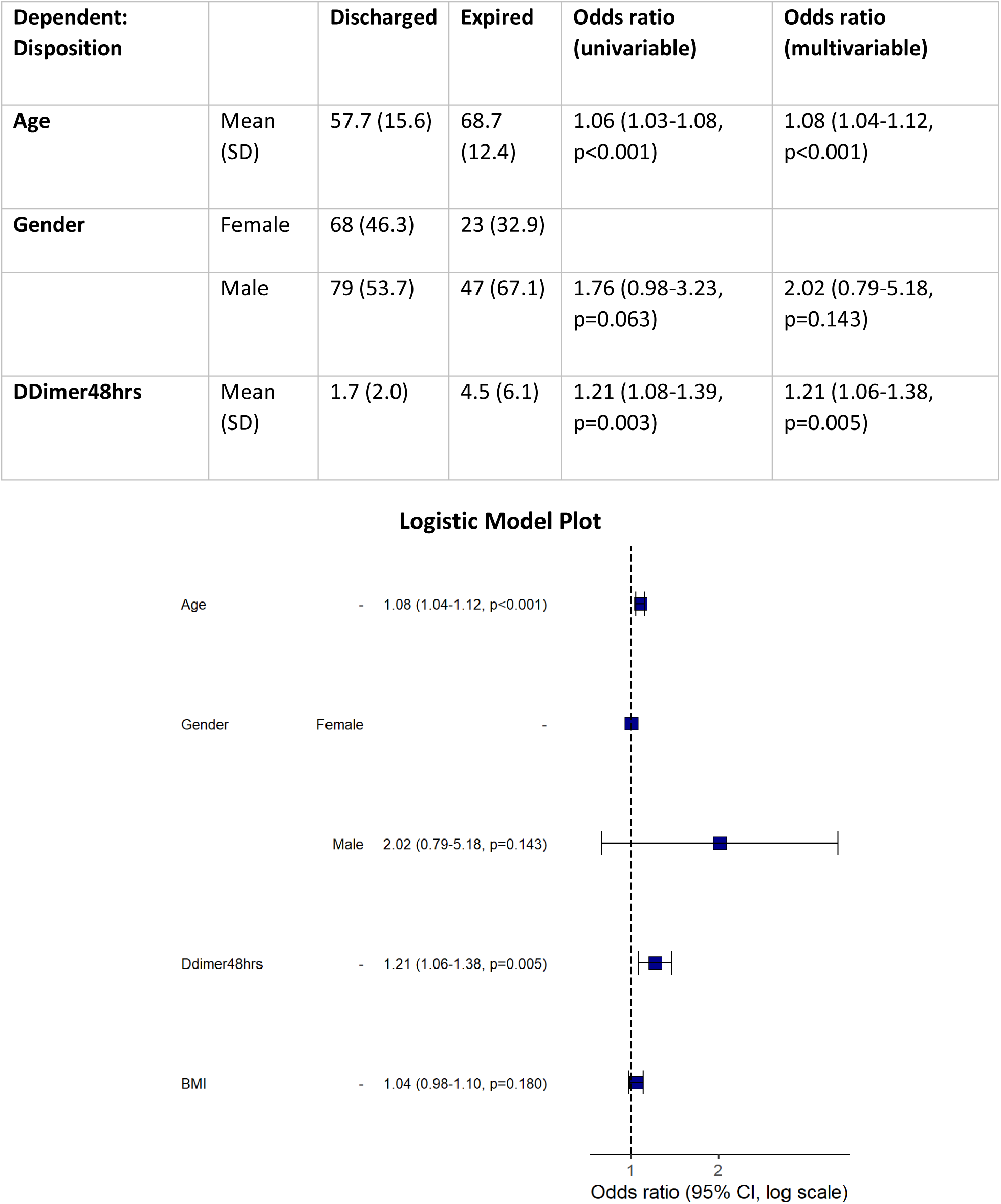
Logistic regression analysis of risk factors for mortality in hospitalized COVID patients. Using univariable and multivariable logistic regression analysis only age and D-Dimer within the first 48 hrs. had significant odds ratio (95% CI, p value).

The receiver operating characteristic curve (ROC) of first 48 hrs. D-Dimer adjusted for age, BMI and gender, showed an area under the curve (AUC) of 0.8 and a very similar AUC with only D-Dimer and age, underscoring D-Dimer as an important admission lab test as predictor of mortality (Figure 2). Using Youden’s J statistic, the optimal cut-point for the initial D-Dimer to predict mortality was found to be 1.65 μg/mL (Figure 3).

**Figure 2.**
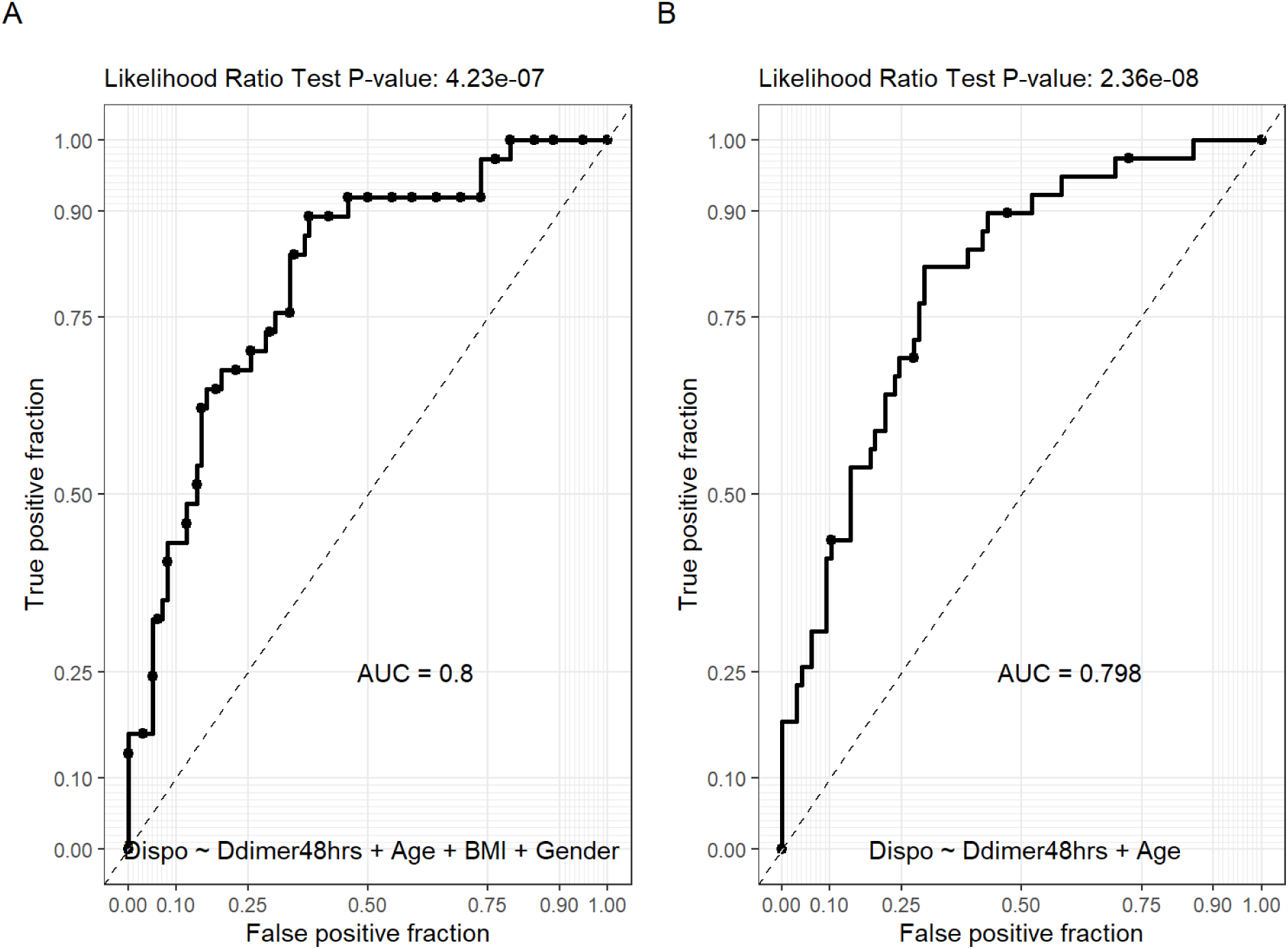
Receiver operating characteristic curve (ROC) of D-Dimer. A. D-Dimer combined with age, BMI and gender, showed an area under the curve (AUC) of 0.8. B. D-Dimer combined with age only displayed an AUC of 0.8.

**Figure 3.**
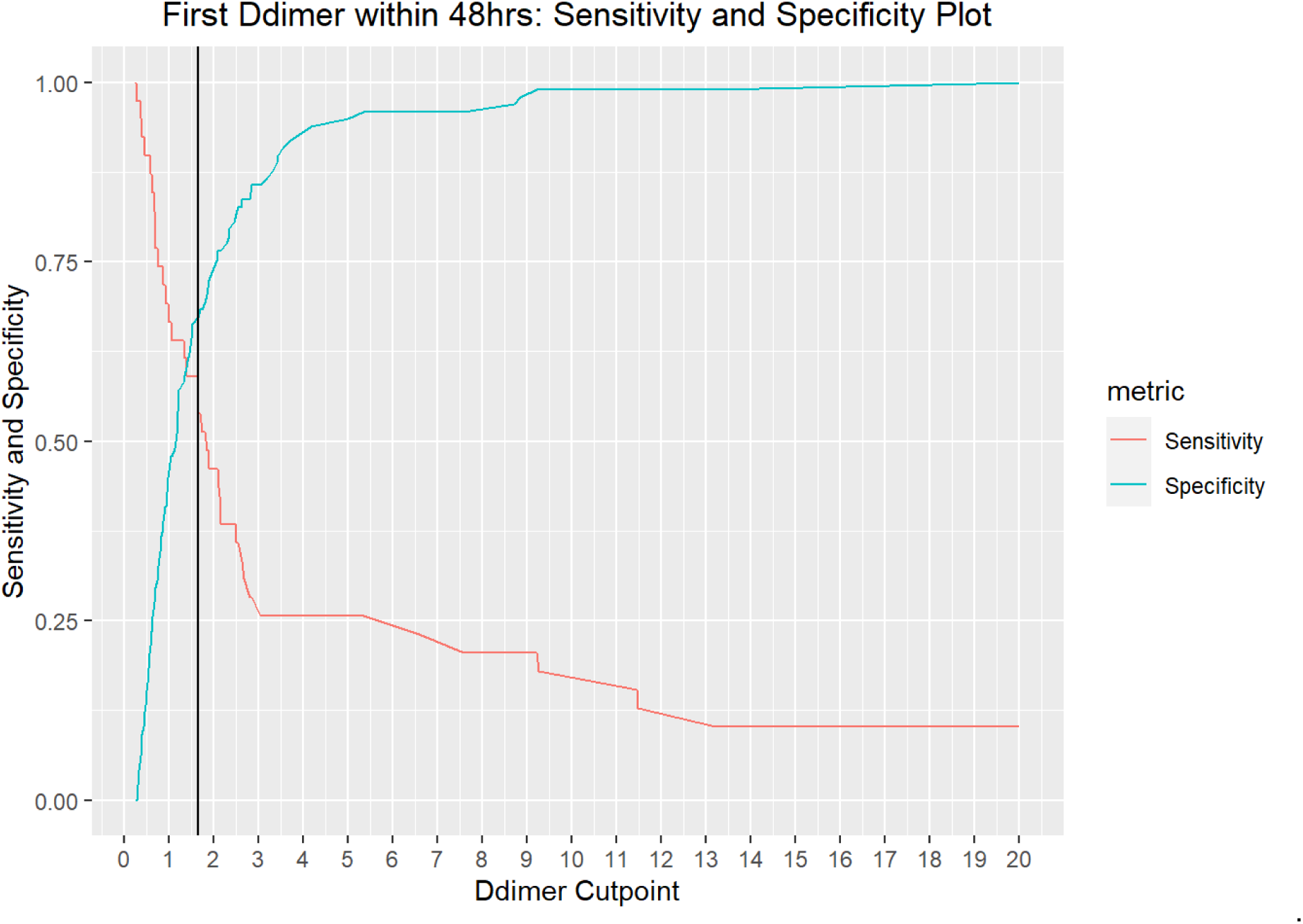
Youden’s J statistic. Youden’s J statistic showed a cut point of D-Dimer 48hrs. to be 1.65 μg/ml. The AUC is 0.63, with a sensitivity of 0.59 and specificity of 0.67.

#### Anticoagulation

178 of the 217 patients were on some anticoagulation. 82.3% (121/147) of those who lived were on anticoagulation as compared to 81.4% (57/70) of those who died; this difference was not significant. Of those who were on anticoagulation, only 12.9% of the survivors or 15.6% of those who survived and were on anticoagulation were given therapeutic doses as compared to 18.6% of those who died. These data, and the medications involved, are detailed in Table 3.

### DISCUSSION

Our cohort consists of 217 patients seen at the main Montefiore Medical Center hospital during the pandemic peak in NYC. Montefiore comprises a population of minorities that is largely underserved and understudied (Black and Hispanics with a minor population of Whites and Asians). The most recent data show that the number of positive cases per 100,000 people was 2,234 – significantly higher than the other boroughs. Survival in our hospitalized patients during this period was poor, with 32% of the hospitalized patients dying. This compares with the death rate of 28.3% for the rest of New York City (7). Given the huge patient load, only the very sick were being admitted in the Bronx so, if true, this may actually compare even more favorably. Although both male gender and black race were increased in those who died vs. those who were discharged, we did not get a significant association. This is in conflict with previous studies which have shown a relationship of black race with increased mortality,(10) but is probably due to the smaller sample size in our cohort which limited comprehensive study of the influences of comorbidities, race and socioeconomic status.

Several published cohorts have examined laboratory characteristics of patient admissions (2, 8, 11). Concordantly with those, we show that D-Dimer within first 48hrs of admission was strongly associated with mortality. Modeling showed that an initial D-Dimer value of 1.65 μg/ml could distinguish between those that would survive and those that would not. Given these data, we would suggest that D-Dimer be factored in the decision-making algorithm of whom to dismiss from the ED. In the Richardson study 2.2% were readmitted within 3 days, although follow-up time was short at 4.5 days (11). Examining the D-Dimer of these patients before discharge may enable us to make more informed decisions. There is anecdotal evidence that thrombotic events may occur in COVID patients post-discharge; following D-Dimers might be a way to distinguish who should get prolonged thromboprophylaxis.

This is the first study of COVID19+ patients in USA that confirms laboratory and clinical observations and the associations with mortality made by Wuhan studies of COVID19 positive patients (2). Goyal et al., described a population in New York City (NYC) that is majority White (37%), minority African Americans (AA) (12.5%) and unknown percentage of Hispanics, as no classification for Hispanics was provided (6). Richardson et al., also described the demographics and comorbidities of a NYC that is majority White (39.8%), followed by Hispanics (23%) and AA (22.6%) (11). Although, Richardson et al., showed laboratory data, no comparison or statistical analysis was shown between discharged patients vs. survivors. In contrast we studied a population of AA (40%), Hispanics (33%) and a minority Whites (18%), representative of the Bronx and overall NYC demographics and we were able to analyze physiological and laboratory parameters as predictors of mortality in our cohort of US COVID19 infected patients.

By analyzing the maximum and minimum levels of important lab parameters we could establish that, in addition to the importance of the initial D-Dimer to screen patients that present with coagulopathy and are at higher risk of mortality, the maximum D-Dimer during hospitalization was also associated with mortality. Huang et., showed that an initial D-Dimer > 1 ug/mL correlates with increased risk of mortality. Our study showed that cut off of 1.65 ug/mL on initial D-Dimer better stratified our patients at higher risk of mortality. No adjustment in D-Dimer for age was done.

It is unclear what the elevation in D-Dimer levels actually signifies: There is evidence of both an increase in venous and arterial disease in COVID and many patients have been demonstrated to have antiphospholipid antibodies (12–14). While we were gathering data, a hospital protocol was developed to place all patients with high D-Dimers on anticoagulation. We do not yet have an idea of the temporal changes in D-Dimer with COVID nor do we know whether these change with anticoagulation. Both are the subject of ongoing research. Our data do not show any differences in outcome but this is from a very small cohort on anticoagulation. Despite many protocols, the effect of anticoagulation on mortality with COVID is uncertain with only a few reports showing that there may be a benefit. Indeed, even the effect of anticoagulation in the symptomatic thromboses/vessel occlusion associated with COVID is unclear (15). It may be that this disease presents with more of a thrombotic microangiopathy (TMA) picture and may be more amenable to TMA therapeutics. Studies are ongoing looking at therapies with anticoagulation, anti-complement, fibrinolysis (16). Should elevations in D-Dimer without evidence of pathology be treated is open to debate. Given the strength of D-Dimer as a predictor of mortality, future studies should focus on establishing guidelines on how to use D-Dimer trending in different settings to better predict mortality, monitor disease progression and response to treatment.

## Data Availability

Data and statistical methods will be available upon request via email

## Acknowledgement

We thank Michael B. Prystowsky, MD, Ph.D (Chair of Department of Pathology/ Montefiore Medical Center The University Hospital of Albert Einstein College of Medicine) for providing invaluable support and resources to all laboratory personnel during the COVID crisis.

We also thank Dr. Eran Bellin, MD (Vice President Clinical IT Research and Development Montefiore Information Technology Professor of Clinical Epidemiology and Population Health Albert Einstein College of Medicine) and Lindsay Stahl for providing guidance about study design and interpretation of results.

